# The Human Leukocyte Antigen Locus and Susceptibility to Rheumatic Heart Disease in South Asians and Europeans

**DOI:** 10.1101/19003160

**Authors:** Kathryn Auckland, Balraj Mittal, Benjamin J Cairns, Naveen Garg, Surendra Kumar, Alexander J Mentzer, Joseph Kado, Mai Ling Perman, Andrew C Steer, Adrian V S Hill, Tom Parks

**Affiliations:** Wellcome Centre for Human Genetics, University of Oxford, Oxford, UK; Babasaheb Bhimrao Ambedkar University, Lucknow, India; MRC Population Health Research Unit, Clinical Trial Service Unit and Epidemiological Studies Unit, Nuffield Department of Population Health, University of Oxford, Oxford, UK; Sanjay Gandhi Postgraduate Institute of Medical Sciences, Lucknow, India; All India Institute of Medical Sciences, New Delhi, India; Fiji National University, Suva, Fiji; Murdoch Children’s Research Institute, Melbourne, Australia; London School of Hygiene & Tropical Medicine, London, UK

**Author notes:** Correspondence to Tom Parks.

**Keywords:** GWAS, HLA, rheumatic heart disease, rheumatic fever, mitral stenosis, *Streptococcus pyogenes*, group A streptococcus

## Abstract

**Background:** Rheumatic heart disease (RHD) remains an important cause of morbidity and mortality globally. Several reports have linked the disease to the human leukocyte antigen (HLA) locus but with negligible consistency.

**Methods:** We undertook a genome-wide association study (GWAS) of susceptibility to RHD in 1163 South Asians (672 cases; 491 controls) recruited in India and Fiji. We analysed directly obtained and imputed genotypes, and followed-up associated loci in 1459 Europeans (150 cases; 1309 controls) from the UK Biobank study. For fine-mapping, we used HLA imputation to define classical alleles and amino acid polymorphisms.

**Results:** A single signal situated in the HLA class III region reached genome-wide significance in the South Asians, and replicated in the Europeans (rs201026476; combined odds ratio 1.81, 95% confidence intervals 1.51-2.18, *P*=3.48×10^−10^). While the signal fine-mapped to specific amino acid polymorphisms within *HLA-DQB1* and *HLA-B*, with conditioning, the lead class III variant remained associated with susceptibility (*P*=3.34×10^−4^), suggesting an independent effect.

**Conclusions:** A complex HLA signal, likely comprising at least two underlying causal variants, strongly associates with susceptibility to RHD in South Asians and Europeans. Crucially, the involvement of the class III region may partly explain the previous inconsistency, while offering important new insight into pathogenesis.

## Background

Rheumatic heart disease (RHD) is the leading cause of cardiovascular death and disability in children and young adults globally [1]. The disease is caused by an aberrant immunological response to *Streptococcus pyogenes* (also termed group A streptococcus), a process that causes scarring and thickening of the heart valves [2]. Beginning in childhood, RHD gradually causes the heart to fail, leading to complications including arrhythmias, stroke and early death [2]. A recent analysis by the Global Burden of Disease Consortium estimated 319,400 deaths and 10.5 million disability-adjusted life-years (DALYs) each year globally due to RHD [3], a substantial disease burden, especially in comparison to other diseases with infectious aetiology [4, 5]. In 2015, the highest age-standardised mortality due to RHD outside Oceania was observed in South Asia, with a total of 119,110 deaths in India alone [3].

Rheumatic heart disease has long been thought to be heritable [6], although until recently, relatively little progress had been made in delineating susceptibility [7]. To date, two genome-wide association studies (GWAS) have been published: the first set in diverse populations in Oceania [8], and the second in Aboriginal Australians [9]. Consistent with several studies predating the GWAS era, which linked the disease to the human leukocyte antigen (HLA) locus on chromosome 6 [10], the Australian study found a signal that peaked in the class II region of HLA just below genome-wide significance, which was fine-mapped to a single nucleotide polymorphism (SNP) located within intron 1 of *HLA-DQA1* [9]. The pre-GWAS results should be interpreted with considerable caution, given the variable genotyping approaches, small sample size, limited quality control and confounding due to genetic ancestry [10]. Nonetheless, it is striking that the specific classical alleles that best explained the Australian signal scarcely feature in the earlier reports, while across the pre-GWAS reports there are no clear examples of the same HLA allele being associated with susceptibility in two or more studies [7, 10].

In contrast to the Australian study, however, our Oceanian study found negligible signal in the HLA locus [8], a surprising finding given the putative role for HLA in the disease’s pathogenesis [11]. While we cannot be certain, it is possible this result represents a false negative, although it is notable the study was adequately powered to detect the large effect sizes that have been reported previously [10]. We speculate that the negative result might be attributable to the substantial genetic heterogeneity within the study population, which could have diluted out a HLA signal, in which the underlying causal variants occurred on distinct background haplotypes in each of the ancestral groups. On balance, while we consider it highly likely that HLA variants contribute to RHD susceptibility, there is a clear need to clarify the causal variants of these association signals.

Accordingly, we undertook a GWAS of susceptibility to RHD limited to the South Asian population, motivated by the need to refine the HLA and other signals and the substantial burden of RHD within this region. We then did a follow-up analysis in individuals of European ancestry in UK Biobank on the basis that robust HLA reference data are available for this very large sample set.

## Methods

### Sample collections

For the South Asian analysis, genetic material was obtained with informed consent from cases and controls recruited to two distinct studies. Specifically, we expanded an existing collection in Northern India [12-16], and we used samples from our existing collection of Pacific Islanders [8], specifically the Fijians of Indian descent. Cases of RHD were defined on the basis of: a history of valve surgery for RHD, a definite RHD diagnosis by echocardiography, or borderline RHD diagnosis by echocardiography with prior acute rheumatic fever [17].

In India, adults with incident or prevalent RHD were recruited as cases from a single large referral hospital, the Sanjay Gandhi Postgraduate Institute of Medical Sciences, Lucknow, Uttar Pradesh; recruitment was limited to patients with an echocardiographic diagnosis of RHD [17]. Controls were recruited based on normal echocardiograms and the absence of prior family history of rheumatic fever [12-16]. In total, DNA samples were obtained from 543 cases and 397 controls. Ethical approval was granted by the Sanjay Gandhi Postgraduate Institute of Medical Sciences (SGPGIMS), as well as the Oxford University Tropical Research Ethics Committee.

In Fiji, children and adults with incident or prevalent RHD were recruited as cases from either the Colonial War Memorial Hospital in Suva, or the Lautoka General Hospital in Lautoka, while members of the general population were recruited as controls, following the approach of the Wellcome Trust Case Control Consortium [18]. Accounting for approximately one third of the population, Fijians of Indian descent are a South Asian population who first came to Fiji from India in the 1870s under the British indentured labour scheme [19]. In total, DNA samples were obtained from 598 cases and 913 controls [8]; of these, 170 cases and 158 controls were of Fijian Indian ancestry. Ethical approval was granted by the Fiji National Health Research Committee and the Fiji National Research Ethics Review Committee, as well as the Oxford University Tropical Research Ethics Committee.

### Array genotyping and quality control

We obtained genetic material by sampling peripheral blood in both Fiji and India. Blood samples collected in India were stored in EDTA and frozen at −20°C until transport to the laboratory facilities at Babasaheb Bhimrao Ambedkar University, Lucknow. Upon arrival, samples were stored at −80°C until extraction using standard salting out procedures. Extracted DNA was prepared for analysis at the Wellcome Centre for Human Genetics (UK), where quantification was performed by PicoGreen (Life Technologies, USA). The handling of blood samples collected in Fiji has previously been described, although it is noteworthy that a proportion of these samples underwent genome-wide amplification due to low DNA concentration [8]. From both collections, 1,268 DNA samples were genotyped at the Oxford Genomics Centre at ∼300,000 variants using the HumanCore-24 BeadChip (Illumina Inc., USA). The resulting data were aligned to the forward strand of the Genome Reference Consortium Human Build 37.

After identifying and removing duplicated variants, the South Asian data was divided into two populations: Fijian Indian (n=328) and Northern Indian (n=940). We employed standard approaches to quality control (QC) the genotyping data [20], with most steps performed using PLINK version 1.9 [21]. Starting with ‘per individual QC’ (Supplementary Figure 1), we measured missingness in each sample and examined its relationship with autosomal heterozygosity. Based on this relationship, we removed individuals if they had a missing data rate ≥3% and if heterozygosity deviated plus or minus three standard deviations from the mean. Due to genotyping batch effects, additional missing data filters were applied to the Northern Indian data: ≥0.3% for batch one, ≥1.1% for batch two and ≥1.6% for batch three (Supplementary Figure 2). In addition, we removed duplicated samples with identity by descent ≥0.9. Finally, by merging the data with the HapMap3 data [22], we identified and removed samples of divergent ancestry.

We then performed ‘per variant QC’. We removed all variants with minor allele frequency (MAF) ≤1.25% because such variants are usually less reliably genotyped [20]. We kept variants with MAF 1.25 to 5% but applied stricter missingness thresholds (Supplementary Figure 1). Finally, we removed variants with deviation from Hardy-Weinberg equilibrium (HWE) *P* <1.0×10^−8^.

### Genome-wide imputation, association testing and meta-analysis

Imputation of genotypes not present on the array or missing was performed using the 1000 Genomes Project phase 3 reference panel [23]. We prephased the variants that had passed QC using SHAPEIT version 2 (r644) [24] before performing genome-wide imputation using IMPUTE2 software [25], excluding imputed SNPs with an information metric ≤0.4, and a MAF ≤5%.

Genome-wide association analysis for the RHD phenotype was performed using a linear mixed model, as implemented in GCTA 1.24.4, which minimises confounding due to population structure, admixture and cryptic relatedness [26]. Additionally, genotypic sex was coded as a covariate for each population, as was sample type (non-amplified or whole genome amplified) for the Fijian Indians and genotyping batch for the Northern Indians. We assessed confounding using quantile-quantile plots and the test statistic inflation factor (λ), and used the accepted threshold for genome-wide significance (*P*<5×10^−8^) [27]. Having estimated effect sizes by transformation [28], we combined the resulting association statistics by genome-wide meta-analysis using inverse-variance-weighted fixed effects, as implemented in METASOFT [29]. Regional association plots, based on those drawn by the widely used LocusZoom software [30], were generated for the data. We collated available data from published GWAS, including the Australian study [9] and a study containing over 200,000 23&Me research participants of European ancestry, of which 1,115 were cases of self-reported rheumatic fever [31].

### HLA imputation analysis

HLA imputation was performed using SNP2HLA [32], a software package that imputes classical HLA alleles and amino acid polymorphisms at class I (*HLA-A, -B* and *-C*) and class II (*-DPA1, -DPB1, -DQA1, - DQB1* and *-DRB1*) loci from SNP data using the Type 1 Diabetes Genetics Consortium (T1DGC) reference panel. The T1DGC reference panel contains 5,868 SNPs and 4-digit classical HLA types for the eight loci listed above for 5,225 unrelated individuals of European ancestry. For comparison, HLA imputation was also performed with the Pan-Asian reference panel (n=530) [33]; this comprises several underlying datasets with ancestry including: Singapore Chinese [34]; Chinese, Indian and Malaysian [34]; and Japanese and Han Chinese from the Phase II HapMap [35]. Association analyses mirrored those for the genotyping data using the imputed dosage data, rather than best-guess genotypes, but excluded alleles or amino acids with imputation accuracy R^2^ ≤0.3.

### Conditional analysis

To identify secondary association signals, conditional association analyses in the SNP-based GWAS and the HLA region were performed with linear mixed models, as implemented in GCTA 1.24.4, using the same covariates as previously mentioned. Within the genome-wide dataset, we first identified the most strongly associated SNP following meta-analysis and performed stepwise iterative conditional regression, adding the dose of the associated SNP as a covariate to the model, to identify other independent signals. We also identified the most strongly associated HLA class I and class II SNPs within this same dataset and performed iterative conditional regression, adding the dose of each associated SNP as a covariate to the model, to identify additional independent signals. Conditional analyses in the HLA region were also performed by adding the dose of each of the previously mentioned SNPs as covariates to the model to see if there were additional signals attributable to HLA alleles or amino acids at each HLA locus.

### Replication analysis

The replication analysis was based on the UK Biobank study, which contains genetic and phenotypic data collected on approximately 500,000 individuals from across the United Kingdom [36]. For the purpose of replication, we used mitral stenosis as a surrogate for RHD. Broadly, with the UK’s low prevalence of RHD, most diagnostic codes indicating RHD will represent other forms of valvular heart disease [37]. In contrast, codes indicating mitral stenosis, which is now a rare finding in the UK population [38], are substantially more likely to indicate underlying RHD [37], as the majority of mitral stenosis cases have underlying rheumatic aetiology [17, 39-42]. Cases were therefore defined by self-report of mitral stenosis at enrolment or an International Statistical Classification of Diseases and Health Related Problems 10^th^ Revision (ICD-10) code for rheumatic mitral stenosis (I05.0, rheumatic mitral stenosis; I05.2, rheumatic mitral stenosis with insufficiency) as a primary or other diagnosis in the hospital episode statistics or on a death certificate. Controls were selected from the remainder of the cohort, matched by age, ethnicity, deprivation index, birth outside the UK and recruitment centre, at a ratio of 1:10, beyond which the performance of linear mixed models deteriorates [43]. In total, we identified 196 cases and 1919 controls, of which 150 cases and 1309 controls were defined as Caucasian (i.e. European) by UK Biobank investigators (Supplementary Table 1). These individuals had previously been genotyped at ∼800,000 variants using the UK Biobank Axiom Array (Affymetrix, USA). These data were quality controlled by removal of individuals with missing rate >2% and variants with missing rate >1%, MAF <5% or HWE *P* <1.0×10^−9^. The remaining preparation of the data, including genome-wide and HLA imputation, and the association analyses, mirrored the process used in the South Asian samples.

## Results

### Genome-wide analysis

In total, 854 Northern Indians (510 cases; 344 controls) and 309 Fijian Indians (162 cases; 147 controls) passed QC and were included (Supplementary Figure 1). A single signal situated in the major histocompatibility complex (MHC) class III region reached genome-wide significance (Supplementary Figure 3A; Supplementary Figure 4A) with minimal evidence of residual confounding (λ=0.9967; Supplementary Figure 3B). The top variant (rs201026476) in this region, with an imputation information metric score of 0.86 for the Fijian Indians and 0.87 for the Northern Indians, had a MAF of 0.15, and each copy of the minor allele was associated with a two-fold increased risk of disease (odds ratio, OR, 1.99, 95% confidence intervals, CI, 1.58-2.51, *P*=7.45×10^−9^). The second and third strongest signals were found in the class I (*HLA-B*, rs3819306, *P*=1.91×10^−7^) and class II (*HLA-DQB1*, rs28724238, *P*=7.77×10^−7^) regions, respectively.

To further define this signal, we performed stepwise conditional analyses by adding the dose of each associated allele as a covariate to the model (Supplementary Figure 4). After conditioning on the class III signal, the strongest signal (rs3819306) was located in *HLA-B* (OR 1.39, 95% CI 1.20-1.61, *P*=1.83×10^−5^; Supplementary Figure 4B). However, conditioning on the lead SNPs in *HLA-B* and *HLA-DQB1*, the lead SNP in class III remained associated with susceptibility (*P*=2.59×10^−4^) suggesting an independent effect (Supplementary Figure 4C). The previously reported rs9272622 [9] was not associated with susceptibility (*P*_LMM_=0.28).

To validate our findings, we examined the HLA locus in the European UK Biobank dataset (150 cases of mitral stenosis; 1309 controls), combining the resulting association statistics with those from our South Asian analyses (Figure 1). The peak SNP in class III was associated with susceptibility in the UK Biobank data in the same direction (rs201026476, *P*_LMM_=0.0057), with a combined effect size that was consistent with the discovery analysis (OR 1.81, 95% CI 1.51-2.18, *P*=3.48×10^−10^; Figure 1A). The variant located in intron 4 of *HLA-DQB1* (rs28724238, OR 1.75, 95% CI 1.42-2.15, *P*=1.73×10^−7^) also replicated (*P*_LMM_=0.017), as did the *HLA-B* signal, although in the combined analysis, the signal peaked at a SNP (rs9405084) located 1,286 base pairs upstream of *HLA-B* (OR 1.36, 95% CI 1.19-1.55, *P*=3.39×10^−6^).

**Figure 1.**
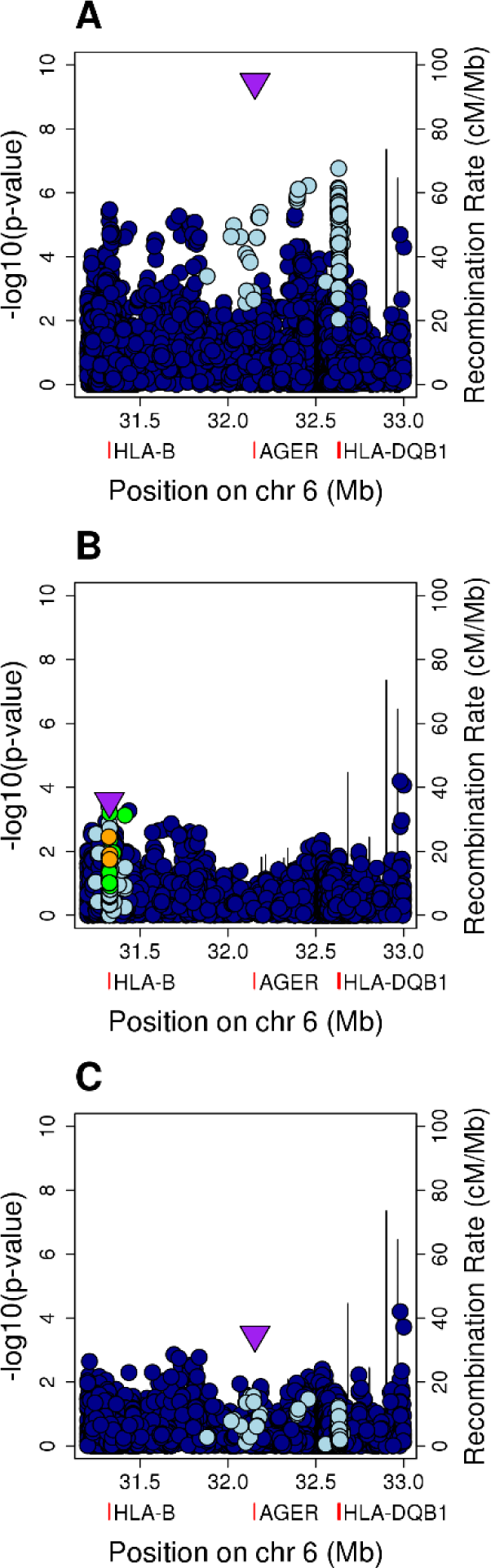
Meta-analysis of the South Asian and UK Biobank data following conditional analyses. **A**, Unconditioned analysis. **B**, Conditioned on the top SNP (rs201026476). **C**, Conditioned on the top class I and class II SNPs (rs9405084 and rs28724238, respectively). For the HLA region, genomic position is plotted against the negative common logarithm of the *P* value from meta-analysis. The top class I (**B**) or class III SNP (**A, C**) following meta-analysis is shown by a purple triangle. Variants are coloured by linkage disequilibrium (LD), with the most associated variant averaged across the entire dataset (estimated r2: dark blue, 0-0.2; light blue, 0.2-0.4; green, 0.4-0.6; orange, 0.6-0.8; red, 0.8-1.0). The location of *HLA-B, HLA-DQB1* and *AGER* are indicated by red rectangles below the x axis. The recombination rate is shown as a line plotted on the right-hand y-axis. These plots are based on those drawn by the widely used LocusZoom software.

The conditional analyses followed a similar pattern, although after conditioning on the top class III SNP, the strongest signal (rs432375, *P*=6.40×10^−5^) was located 2,290 base pairs upstream of *HLA-DOA*, a HLA class II alpha chain paralogue, rather than at *HLA-B* (Figure 1B). However, the class I signal remained apparent, with the lead SNP a coding variant within exon 1 of *HLA-B* (rs1050462, *P*=2.69×10^−4^). After conditioning on both rs9405084 (class I) and rs28724238 (class II), the class III signal was again maintained (rs201026476, *P*=3.34×10^−4^; Figure 1C).

### HLA imputation analysis

To further understand the potential functional variants across the HLA region, we imputed classical HLA alleles and amino acid polymorphisms at class I and class II loci. Using the T1DGC reference panel, a reasonably high proportion of variants were accurately imputed based on the R^2^ metric (proportion variants with R^2^ >0.80: Fijian Indian, 91.74%; Northern Indian, 91.52%; European UK Biobank, 96.16%). For comparison, when using the Pan-Asian reference panel, imputation accuracy was significantly lower (proportion variants with R^2^ >0.80: Fijian Indian, 73.10%; Northern Indian, 71.10%).

The strongest allelic signal in the class II region in the South Asian analysis mapped to the *HLA-DQB1*\*03:03 allele (OR 1.90, 95% CI 1.41-2.55, *P*=2.59×10^−5^; Supplementary Figure 4A; Supplementary Figure 5A), an allele imputed with high accuracy (Supplementary Table 2). While this signal was maintained in the combined European and South Asian analysis (OR 1.78, 95% CI 1.38-2.29, *P*=1.00×10^−5^; Figure 2A; Supplementary Figure 6A), it was weaker than that at the coding change at position 185 (Thr185Ile; rs1130399) of *HLA-DQB1* (Figure 3A) which was associated with a 1.5-fold increased risk of disease (OR 1.56, 95% CI 1.31-1.85, *P*=3.95×10^−7^; Figure 3B). There was also a signal at *HLA-B*\*40:06 (*P*=4.82×10^−4^; Figure 2A), although again the signal was slightly stronger at the coding change at position −16 (Val-16Leu; rs1050462) of *HLA-B* (*P*=5.67×10^−5^; Figure 3A; Supplementary Figure 6C).

**Figure 2.**
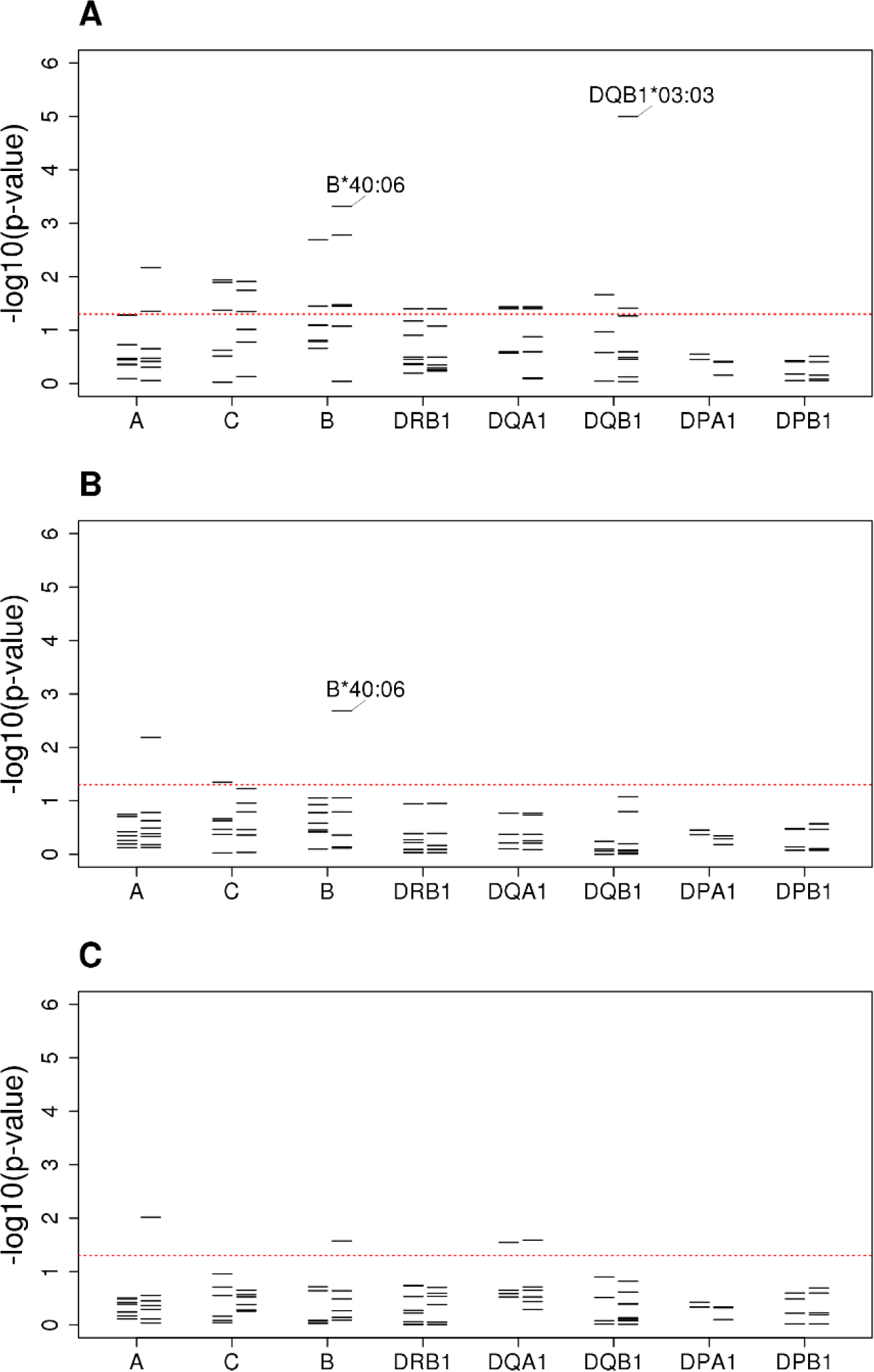
Classical HLA alleles associated with susceptibility to RHD within the South Asian and UK Biobank data following conditional analyses. **A**, Unconditioned analysis. **B**, Conditioned on the top SNP (rs201026476). **C**, Conditioned on the top class I and class II SNPs (rs9405084 and rs28724238, respectively). For each locus, the negative common logarithm of the *P* value from LMM analysis is plotted with two-digit alleles to the left and four-digit alleles to the right defined by HLA imputation using SNP2HLA software with the T1DGC reference panel.

**Figure 3.**
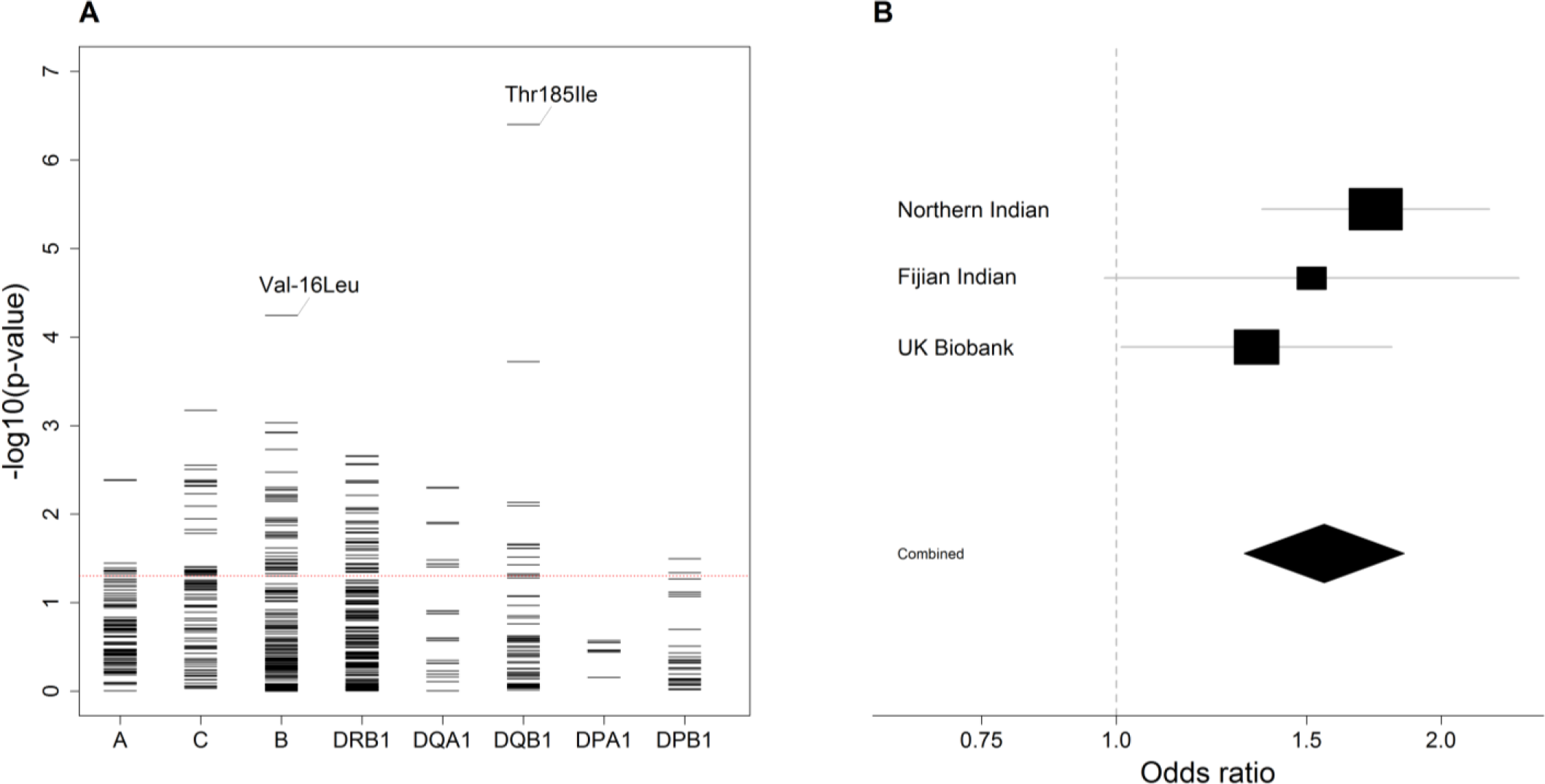
Amino acid variants following HLA imputation. **A**, For each locus, the negative common logarithm of the *P* value from LMM analysis is plotted for each amino acid polymorphism defined by HLA imputation. For *HLA-DQB1* Thr185Ile and *HLA-B* Val-16Leu, the effect is shown in a single direction only. **B**, Forest plot for the presence of isoleucine at position 185 in *HLA-DQB1*. For each population, the black squares centre on the odds ratio estimate from LMM on a logarithmic scale; the size of the square is proportional to the weight of the analysis. The horizontal line through each square corresponds to the confidence intervals. The black diamond centres on the combined effect estimate by fixed effects meta-analysis and stretches to the confidence intervals; the dashed line indicates no effect.

Overall, there was limited signal at the classical alleles and amino acids linked to susceptibility in the Australian study mentioned above, although we did observe an effect at the coding change at position 38 of *HLA-DQB1* in the same direction (OR 0.87, 95% CI 0.76-0.99, *P*=0.031; Supplementary Table 3). Interestingly, however, *HLA-DQB1*\*03:03 was the classical allele, with MAF >0.5%, most associated with a self-reported history of rheumatic fever risk in a study by 23&Me (OR 1.28, 95% CI 1.05-1.55, *P*=0.017) [31], with an effect consistent in size and direction (combined OR 1.45, 95% CI 1.24-1.69, *P*=4.05×10^−6^; Supplementary Figure 6B).

Finally, as in the SNP-based GWAS, the signal in class II was linked to the class III signal, while the class I signal was independent. After conditioning on the class III SNP, the strongest signal was *HLA-B*\*40:06 (*P*=0.0021; Figure 2B). Conditioning on both the class I and class II SNPs, only a marginal signal remained at *HLA-A*\*02:11 (*P*=0.0096; Figure 2C).

## Discussion

In the first genome-wide association study of RHD to be reported outside of the Australia-Pacific region, we have resolved a complex HLA signal into its component parts. We have shown that a single HLA signal overlapping the class III region most likely comprises at least two independent coding or regulatory effects across the class I, II and III loci. While most studies to date have focused on the relationship between classical HLA alleles and susceptibility, our data suggest these signals are in fact more complex and cannot be attributed to the classical alleles alone. Indeed, based on annotations in Ensembl [44], the effect of Thr185Ile in *HLA-DQB1*, as an example, is much more likely to be regulatory than coding, not least because it shows a strong negative association with expression of *HLA-DQB1* itself [44]. Similarly, the independent lead class III variant (rs201026476), situated in the 3 prime UTR of the *PBX2* (Pre-B-cell leukaemia transcription factor 2) gene has regulatory annotations and thus could impact expression of one or more of the numerous immunologic genes in the class III region.

While the role of HLA polymorphism has long been suspected, there remains some doubt about the roles that individual alleles play in disease susceptibility across populations. Importantly, our analysis represents the first time HLA signals for RHD have been demonstrated with consistent direction and effect size in more than one ancestral group. Moreover, the signal at *HLA-DQB1*\*03:03 in the 23&Me study, although based on self-reported rheumatic fever [31], adds further weight to our findings. That our results differ from those reported in the Australian study [9] is unsurprising, given there are likely to be substantial differences between the HLA loci of South Asians and Aboriginal Australians. Added to this, there were also a number of methodological differences, including the software employed for HLA imputation and linear mixed model analysis, which may exacerbate any disparity. Nonetheless, it is reassuring that both studies observed a signal at the coding change at position 38 of *HLA-DQB1*, raising the possibility that the two studies are tagging the same underlying causal variants. As noted above, we observed negligible HLA signal in our study set in Oceania including, beyond the Fijian Indian subgroup, the specific variants that associate with susceptibility in this South Asian analysis. Indeed, it may be difficult to fully unravel the contribution of HLA to RHD susceptibility in individuals of Oceanian ancestry until further HLA data are generated from these populations, enabling HLA imputation with a population-specific reference panel. Accordingly, we have begun efforts to develop such a panel by HLA typing a subset of our samples from individuals with Oceanian ancestry. Relating our findings to the HLA signals reported before the GWAS era is more difficult, not least because of the marked inconsistencies and the limitations of the studies themselves in addition to true geographical and ancestral differences. Interestingly, the presence of a signal in the class III region, which could have been differentially tagged in earlier studies, goes some way to explaining the inconsistencies of previously reported HLA associations.

This study has a few limitations. First, in comparison to some contemporary GWAS, our total sample size is relatively modest, and hence it is likely many variants with smaller effects will go undetected until larger collections are assembled. Nonetheless, our study was well powered to detect the vast majority of large effect variants reported in the candidate gene era [10]. Second, within Fiji, members of the general population were recruited as controls; these individuals did not undergo echocardiograms and therefore it is possible to have included a small number of undiagnosed cases of RHD. However, the prevalence of definite RHD among Fijians of Indian descent has been estimated at 3.6-4.4 cases per 1,000 [45, 46] such that the impact of misclassification should be minimal. There may also be shortcomings associated with using the UK Biobank study, for there were no echocardiographic diagnoses available. However, specificity is likely to be regained by limiting the analysis to the mitral stenosis subgroup, an approach that is somewhat validated by the consistent replication of the South Asian signals.

Third, the genotyping array, containing ∼230,000 variants following QC, was not very dense and contained, in comparison to other genotyping arrays, a limited number of variants within the HLA region. Despite this, overall HLA imputation accuracy was high when using the T1DGC reference panel. Imputation accuracy is highly dependent upon the reference panel used and as such, we have so far deliberately limited these analyses to the South Asians and Europeans for whom there are reasonable reference panels available. Fourth, this report is focused on the HLA locus because it was the only region of the genome that reached genome-wide significance in the South Asian analysis. Efforts are underway to combine these and other datasets in a genome-wide meta-analysis, facilitating follow-up of other regions, such as the immunoglobulin heavy chain locus [8]. Finally, at this stage, we cannot resolve the genetic determinants of sub-phenotypes, such as specific valve lesions, disease progression or complications, these are issues which larger-scale collaborative datasets should begin to tackle.

In summary, we report a major susceptibility locus for RHD in the HLA region, likely comprising at least two underlying causal variants which strongly associate with susceptibility to RHD in South Asians and Europeans. These findings add substantially to the knowledge of the role of HLA polymorphism in susceptibility to this devastating and neglected disease. This not only has important ramifications for understanding the immunogenetic basis of the disease process, but also offers important new insight into pathogenesis.

## Data Availability

Genotype and phenotype data underlying the manuscript have been deposited in the European Genome-phenome Archive under accession numbers EGAS00001001881 and EGAS00001003565.

## Data availability

Genotype and phenotype data underlying the manuscript have been deposited in the European Genome-phenome Archive under accession numbers EGAS00001001881 (Fijian Indian data) and EGAS00001003565 (Northern Indian data). Some restrictions on access and usage apply and use is restricted to research focused on RHD.

## Supplementary data

Supplementary materials are available at XXXX.

## Author contributions

B.M., A.C.S., A.V.S.H. and T.P. organised and designed the study. B.M., A.C.S., A.V.S.H. and T.P. managed the study. B.M., N.G., S.K., J.K., M.L.P. and T.P. recruited the patients. K.A. and S.K. did the laboratory studies. K.A., A.J.M. and T.P. did the statistics and bioinformatics. K.A., B.M., B.J.C., A.J.M., A.V.S.H. and T.P. contributed to the interpretation of the results. K.A. wrote the first draft of the manuscript under the supervision of T.P. All authors contributed to revisions and approved the final version for publication.

## Acknowledgements

We thank the investigators of the Pacific Islands Rheumatic Heart Disease Genetics Network which facilitated the sample collection in Fiji. We also thank the UK Biobank Resource (application number 11537), on which part of this research has been conducted.

